# Comparing dynamics and determinants of SARS-CoV-2 transmissions among health care workers of adult and pediatric settings in central Paris

**DOI:** 10.1101/2020.05.19.20106427

**Authors:** A Contejean, J Leporrier, E Canouï, F Alby-Laurent, E Lafont, L Beaudeau, P Parize, F Lecieux, A Greffet, G Chéron, R Gauzit, J Fourgeaud, AS L’Honneur, JM Tréluyer, C Charlier, A Casetta, P Frange, M Leruez-Ville, F Rozenberg, O Lortholary, S Kernéis

## Abstract

**Background:** From the start of the pandemic, health-care workers (HCW) have paid a heavy toll to the coronavirus disease-19 (COVID-19) outbreak.

**Objectives:** To describe the dynamics and determinants of severe acute respiratory syndrome coronavirus-2 (SARS-CoV-2) infection in HCW.

**Design:** Prospective observational study conducted from February 24^th^ until April 10^th^, 2020.

**Setting:** Comparison of a 1,500-bed adult and a 600-bed pediatric setting of a tertiary-care university hospital located in central Paris.

**Participants:** All symptomatic HCW screened for SARS-CoV-2 on a nasopharyngeal swab.

**Measurements:** HCW screened positive were prospectively questioned on their profession, symptoms, occupational and non-occupational exposures to SARS-CoV-2.

**Results:** Among 1344 symptomatic HCW tested, 373 were positive (28%) and 336 (90%) corresponding questionnaires were completed. Three hospitalizations and no death were reported. Most HCW (70%) had patient-facing occupational activities (22% in COVID-19 dedicated units). The total number of HCW cases peaked on March 23^rd^, then decreased slowly, concomitantly with a continuous increase of compliance to preventive measures (including universal medical masking and personal protective equipment (PPE) for direct care to COVID-19 patients). Attack rates were of 3.2% and 2.3% in the adult and pediatric setting, respectively (p=0.0022). In the adult setting, HCW more frequently reported exposure to COVID-19 patients without PPE (25% versus 15%, p=0.046). Report of contacts with children attending out-of-home care facilities dramatically decreased over the study period.

**Limitations:** Lack of COVID-19 negative controls and recall bias.

**Conclusion:** Universal masking, reinforcement of hand hygiene, and PPE with medical masks for patients’ care allowed protection of HCW and containment of the outbreak. Residual transmissions were related to persistent exposures with undiagnosed patients or colleagues and not to contacts with children attending out-of-home care facilities.

## Introduction

From the very start of the pandemic, health-care workers (HCW) have been particularly exposed to nosocomial transmissions of severe acute respiratory syndrome coronavirus-2 (SARS-CoV-2). As of February 11^th^, 2020, China reported more than 1,700 infected HCW in Hubei alone (1), contributing to 3.8% of total coronavirus disease-19 (COVID-19) cases, and at least 23 had died (2). Occupational transmission of SARS-CoV-2 to HCW was lately reported in other countries as in the United Kingdom (UK)(3,4) and the United States (US) (5).

Front-line HCW already paid a heavy price to previous Coronavirus outbreaks. During the 2003 SARS epidemic in Singapore, the index patient started off a chain of nosocomial cases resulting in transmission to 60 HCW, with attack rates of up to 32% for ward-based staff (6). Thus, nosocomial transmissions have been recognized as an important amplifier in Coronavirus epidemics. Cross-transmissions both drive a shortage of HCWs who are isolated once infected and raise anxiety and fear among hospital staff. This cascading effect further contributes to saturation of the health care system.

Protection of HCW is therefore a key concern. As the pandemic declared then accelerated, knowledge on ways of transmission of SARS-CoV-2 also advanced. Epidemiological data and temporal patterns of viral shedding now suggest that infectiousness starts from two days before symptom onset and declines quickly within seven days (7), at least in immunocompetent individuals. The virus mainly spreads by droplets transmission and may survive on surfaces up to 72 hours (8). Most national and international guidelines recommend that HCW in contact with COVID-19 patients should wear personal protective equipment (PPE) that includes gowns, gloves, eye protections, medical masks for standard care and FFP2 during aerosol-generating procedures (9–13).

In France, the first imported case of COVID-19 was detected on January 24th, 2020 in a 31-year-old Chinese male tourist from Wuhan (14). By February 29^th^, 2020, a month since the notification of the first case, a total of 100 COVID-19 cases had been confirmed in France (https://www.santepubliquefrance.fr). Two administrative regions (Ile-de-France, including Paris, and Grand-Est) were most rapidly and severely affected. Social distancing strategies were successively implemented by the French Government by mid-March 2020. In health-care settings, in addition to PPE for direct care for COVID patients, universal masking was recommended for all hospital employees.

Symptomatic staff were recommended systematic screening for SARS-CoV-2 and prompted to stay isolated at home for seven days when proven positive. Nevertheless, as of April 20^th^, 4,180 professionals were infected in the hospitals of the Assistance Publique-Hôpitaux de Paris (AP-HP, the largest French hospital institution, accounting for about 100,000 employees [http://www.aphp.fr]). Questions raised on the routes of transmission explaining these persistent contaminations and on the respective role of in-hospital and out-hospital exposures. Indeed, targeting effective strategies to curtail the epidemic among HCW relies on a preliminary assessment of residual sources of transmissions. Here, we describe the spread of the COVID-19 outbreak among HCW of two settings of the AP-HP (one mainly caring for children and the other adults) and compared their occupational and non-occupational exposures to SARS-CoV-2.

## Material and method

### Setting

The study was conducted in two settings of the 2,100-bed tertiary-care university hospital (AP-HP. Centre, Université de Paris) located in central Paris, France. Cochin-Broca (“Adult setting”) is a 1,500-bed healthcare setting that includes acute care, rehabilitation and long-term units mainly caring for adult patients, except for a neonatology unit (63 beds) linked to the obstetrical ward. Necker (“Children setting”) is mainly dedicated to children care (436 beds), and additionally includes five medical wards (150 beds) caring for immunocompromised adults (hematology, infectious diseases and nephrology-kidney-transplantation) along with an obstetrical unit and an intensive care unit (ICU). Both settings include medical, surgical, obstetrical and intensive care units qualified to provide care to COVID-19 patients. As of March 10^th^, 2020, 7,916 and 5,362 employees were regularly working in the Adult and Children setting respectively. Triage and management of COVID-19 patients were comparable in both settings. All patients with either clinical and/or thoracic computed tomography findings consistent with a COVID19 infection were screened on a nasopharyngeal swab or a respiratory invasive sample on admission. Patients screened positive were referred to dedicated wards with dedicated trained personnel.

### Participants and interventions

Social distancing strategies were successively implemented by the French Government on March 12^th^ (school closure) and March 17^th^ (widespread closures, restriction of business and transport). Of note, schools and nurseries remained opened for children of hospital staff all along the epidemic period.

In health-care settings, from the start of the epidemic in February 2020, PPE was recommended for HCW caring for suspected or confirmed COVID-19 patients. In brief, PPE consisted in gowns, gloves, eye protections and either medical masks for standard care or FFP2 masks during airway aerosol-generating procedures. Universal masking with medical masks for all hospital employees was advised from March 16^th^ in our institutions. Testing for SARS-CoV-2 of symptomatic staff started on February 24^th^ in the Adult setting, and on March 5^th^ in the children setting. In both settings, hospital employees presenting either with fever (reported or measured >37.8°C), cough, rhinorrhea, muscle pain, shivers, loss of smell or taste, unusual persistent headaches or severe asthenia, were referred to the two on-site screening pods. Trained medical doctors or nurses collected a nasopharyngeal swab for each symptomatic staff member. Test results were communicated within 24 hours via a secured email or by phone. According to the institution policy, staff with positive results were sent home and able to return to work after seven days (including two days after resolution of any symptoms) if they felt well enough to do so.

### Data

Shortly thereafter, HCW (all hospital employees) with positive results were prospectively contacted by phone by the clinical research team and invited to participate to the study. After three unsuccessful attempts, the participant was considered unreachable. During a phone interview, the following data were collected on a standardized questionnaire: age, gender, profession, symptoms and date of onset, and exposure to SARS-CoV-2 within the 10 days preceding symptoms onset. The study was basically designed to assess sources of exposures of HCW and for confidentiality reasons, we did not collect precise data on past medical history and comorbidities. Exposures were classified as: in-hospital related to patients’ care (average number of close contacts per day with COVID-19 patients with and without PPE, compliance to infection prevention and control [IPC] protocols), in-hospital related to other activities (contacts with colleagues during meal breaks, meetings etc.) and out-hospital (frequentation of public transports, contacts with household members, especially children kept outside the household). A contact at a distance <2 meters for >10 minutes was defined as close contact (14).

### Virology methods

SARS-CoV-2 was detected in nasopharyngeal samples by amplification of E, RdRp and N genes using the Allplex Eurobio® reagent as recommended by the manufacturer or the RealStar® rtPCR kit, a triplex PCR amplifying the viral genome in the E and S genes and an internal control. The result was considered positive if three out of the three targets were amplified. A control sample was requested if only one or two target genes were amplified.

### Statistical methods

The number of cases was computed on a daily basis. Continuous variables are presented as medians (interquartile ranges, IQR) and categorical variables as numbers (percentages). Fischer’s exact tests were used for comparisons of qualitative variables and Mann Whitney’s tests for quantitative variables. All tests were two-sided with a 0.05 value for significance. For better clarity, mobile means were calculated and displayed on the graphs. Statistical analyses were performed using the R software (3.3.2, R Foundation for Statistical Computing, Vienna, Austria).

### Ethics

This study was approved by the Ethical Review Committee for publications of the Cochin university Hospital (CLEP) (N°: AAA-2020-08012). According to French policy, a non-opposition statement was obtained for all participants, meaning that all had received written detailed information on the objectives of the study and were free to request withdrawal of their consent for participation at any time.

### Role of the funding source

This study had no funding source or sponsor implicated in the study design, in the collection, analysis, and interpretation of data, in the writing of the report, and in the decision to submit the article for publication.

## Results

As early as from February 24^th^ to April 10^th^, 1,344 symptomatic HCW were tested for SARS-CoV-2 over a total of 13,278 employees in both settings (10%, Adult setting [866/7916, 10.9%], Children setting [478/5,362, 8.9%]). Overall, 373/1,344 (28%) tested positive (Adult setting [251/866, 29%], Children setting [122/478, 26%]), leading to an overall attack rate of 2.8% (Adult setting [251/7,916, 3.2%], Children setting [122/5,362, 2.3%] p=0.0022). Figure 1 details daily breakdown of SARS-CoV-2 testing in the Adult setting (Figure 1A) and in the pediatric setting (Figure 1B). The total number of cases peaked on March 23^rd^, 2020 then decreased slowly until April 10^th^. Patterns were different between the two settings, and the outbreak appeared to be more intense in the Adult setting, particularly in the early phase of the epidemic, with a more rapid increase of number of infected HCW. A residual number of cases was observed in both settings after the peak with similar paces. Figure 1C illustrates the crude number of daily consultations in emergency departments for suspected or confirmed COVID-19 in Paris during the study period. The epidemic peaked in the general population in Paris on March 31^st^.

**Figure 1:**
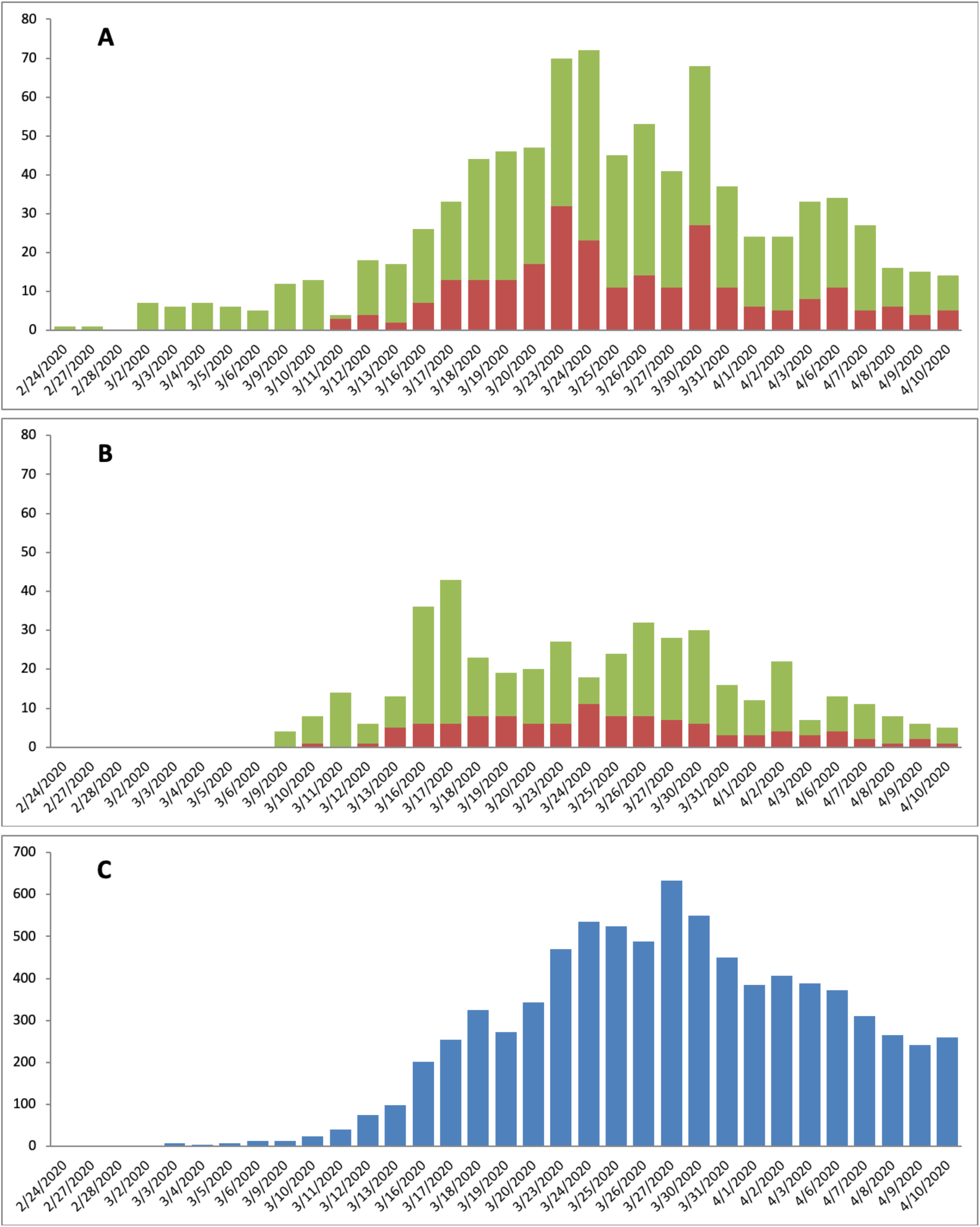
Epidemic curves. A: epidemic curve in the Adult setting. B: epidemic curve in the Children setting. C: Number of daily hospitalizations for COVID-19 in the hospitals of Paris area. Red bars represent the COVID-19 positive cases among hospital staff; green bars represent the hospital personnel tested negative for COVID-19; blue bars represent the crude number of daily consultations in emergency departments for suspected or confirmed COVID-19 in central Paris (data provided by https://geodes.santepubliquefrance.fr/.

Overall, 336/373 hospital personnel (90%) answered to the phone interview, two refused and 35 were unreachable. Main characteristics of the participating HCW and their symptoms are presented in Table 1. In both centers, the large majority were women (265/336, 79%), with direct patient-facing activities (234/336, 70%). Most were posted outside COVID-19 dedicated wards (261/336, 78%). Employees were younger in the pediatric setting. The most frequently reported symptoms were asthenia and headaches and 118/336 (35%) did not report any fever or had a body temperature below 38°C. Patients reporting measured or subjective fever had a median temperature of 38.5°C (minimal 36.2°C, maximal 41.0°C). Three staff members were hospitalized. No death was reported.

**Table 1:** Study population characteristics. Results are presented as n (%) or median [Q1-Q3]

|  | **Overall cohort**<br>**(n=336)** | **Adult setting**<br>**(n=227)** | **Children setting**<br>**(n=109)** | **p-value** |
| --- | --- | --- | --- | --- |
| **Age, years** | 40 [30-53] | 42 [31-53] | 37 [29-50] | < 0.0001 |
| **Sex (female)** | 265 (79) | 180 (79) | 85 (78) | 0.78 |
| **Professional category** |  |  |  |  |
| Physicians | 91 (27) | 64 (28) | 27 (25) | 0.21 |
| Paramedic staff (nurses, care assistants...) | 205 (61) | 132 (58) | 73 (67) |  |
| Administrative staff | 28 (8) | 20 (9) | 8 (7) |  |
| Other employees (kitchen, child carriers...) | 12 (4) | 11 (5) | 1 (1) |  |
| **Occupational activities with direct patient facing** | 234 (70) | 161 (71) | 73 (67) | 0.53 |
| **Symptoms** |  |  |  | 0.17 |
| Asthenia | 272 (81) | 176 (78) | 96 (88) |  |
| Headaches | 262 (78) | 172 (76) | 90 (83) |  |
| Fever (measured or reported) | 246 (73) | 169 (74) | 77 (71) |  |
| Cough | 227 (68) | 143 (63) | 85 (77) |  |
| Anosmia | 229 (68) | 146 (64) | 83 (76) |  |
| Muscle pain | 220 (66) | 145 (64) | 75 (69) |  |
| Ageusia | 215 (64) | 134 (59) | 81 (74) |  |
| Rhinorrhea | 171 (51) | 118 (52) | 51 (49) |  |
| Diarrhea | 129 (38) | 88 (39) | 41 (37) |  |
| Dyspnea | 126 (37) | 80 (33) | 46 (42) |  |
| **Median time from symptom onset to screening** | 3 [1-4] | 3 [1-4] | 2 [1-4] | 0.59 |

Exposures within 10 days prior to onset of symptoms reported by participants are presented in Table 2. The majority recalled a contact without PPE with an index case. Iterative contacts with patients without PPE were more frequently reported by hospital staff from the Adult setting (56/227 [25%] versus 16/109 [15%] in Children setting, p=0.046), whereas contact with index cases in the household or with colleagues were more frequent in the Children setting. Most employees declared wearing a mask always/most of the time at hospital, but 65/336 (19%) admitted taking off masks during breaks in the presence of other colleagues (204/336, 61% during lunch breaks). More than half (201/336, 60%) reported using public transportation but less than 25% (82/334) wore mask outside home. Mean time spent in public transports was over one hour per day in 56% (112/201). The large majority reported performing hand hygiene systematically when returning home (303/336, 90%). Forty-seven (14%) had children aged 0-4 years and 78 (23%) aged 5-15 years in the household. Sixty (18%) reported having children kept by child careers outside the family home: 42 at school (70%), 18 at nursery (30%), and 9 (15%) by other persons (grandparents, neighbors). The large majority (54/60, 92%), were in childcare facilities welcoming more than five children simultaneously. Although not precisely assessed, several cases were retrospectively linked to clusters of contaminations identified at the Ile de France regional level among HCW who attended social events that took place before implementation of social distancing measures.

**Table 2:** Exposures reported by 336 HCW tested positive for SARS Cov2, in the 10 days preceding symptoms onset. Results are presented as n (%). * distance <2 meters for >10 minutes; ^†^PPE: Personal Protective Equipment

|  | Adult setting<br>(n=227) | Children setting<br>(n=109) | p-value |
| --- | --- | --- | --- |
| **At least one close contact\* with an index case without PPE†** | 141 (62) | 76 (70) |  |
| Patient | 57 (25) | 23 (21) | 0.49 |
| Household member | 35 (15) | 31 (28) | 0.0079 |
| Children <15 years | 7 (3) | 9 (8) |  |
| Colleague | 81 (36) | 52 (49) | 0.043 |
| **Exposure to patients** |  |  |  |
| Regularly posted in a unit caring for COVID19-patients | 50 (22) | 25 (23) | 0.89 |
| Has ≥1/day close contact with suspected or confirmed COVID19 patients without PPE† | 56 (25) | 16 (15) | 0.046 |
| **Exposure to colleagues** |  |  |  |
| Wears a medical mask always/most of the time at hospital | 141 (63) | 68 (64) | 0.90 |
| Spends on average >1 hour/day with colleagues without mask | 78 (35) | 33 (33) | 0.80 |
| Has on average >4 close contacts/day with colleagues without mask | 118 (52) | 63 (59) | 0.28 |
| **Out-of-hospital exposure** |  |  |  |
| Uses public transports | 134 (59) | 67 (62) | 0.72 |
| Wears a mask always/most of the time outside home | 61 (27) | 21 (19) | 0.14 |
| Leaves home on average more than once a week | 191 (84) | 99 (91) | 0.13 |
| Lives with ≥ 2 additional household members | 117 (52) | 55 (51) | 0.91 |
| Has ≥1 child kept outside the household (school, nursery) | 38 (17) | 22 (21) | 0.36 |

We carried out a second analysis, focusing on HCW with direct patient facing activities (n=234), comparing those caring for adults to those caring for pediatric patients, regardless of the geographic location. Overall, 169 HCW were posted in adult units (either from the Adult setting [n=160] or from the Children setting [n=9]) and 65 in pediatric units (1 from the neonatology ward of the Adult setting and 64 from various medical and surgical wards of the Children setting). Results are detailed in the Supplementary Appendix. The proportion of HCW reporting contacts with COVID-19 patients without PPE was even higher (35% in adult staff versus 13% in pediatric staff, p=0.00060).

Figure 2 presents the dynamics of various exposures reported by positive hospital personnel over time. Exposure to COVID-19 patients with PPE increased (Figure 2A), in parallel to the rise of COVID-19 patients’ admissions. The rate of HCW reporting contact with COVID-19 patients without PPE was relatively stable around 20% over the study period (Figure 2B). The number of patients’ admissions was higher in the Adult setting, consistent with the number of cases reported in adults compared to children in the general French population (Supplementary Figure). Concomitantly, compliance with mask wearing increased (Figure 2 C-D), but at the end of the study period, a residual proportion of staff still reported contacts without PPE with COVID-19 patients and with colleagues without masks. This was more marked in the Adult setting, where between April 3-10, eight personnel reported contacts with COVID-19 patients, including four in the same geriatric ward. Mask wearing outside home increased but capped around 60% as of April 10^th^ (Figure 2E). Conversely, the proportion reporting childcare outside home fell dramatically over the study period (Figure 2F).

**Figure 2:**
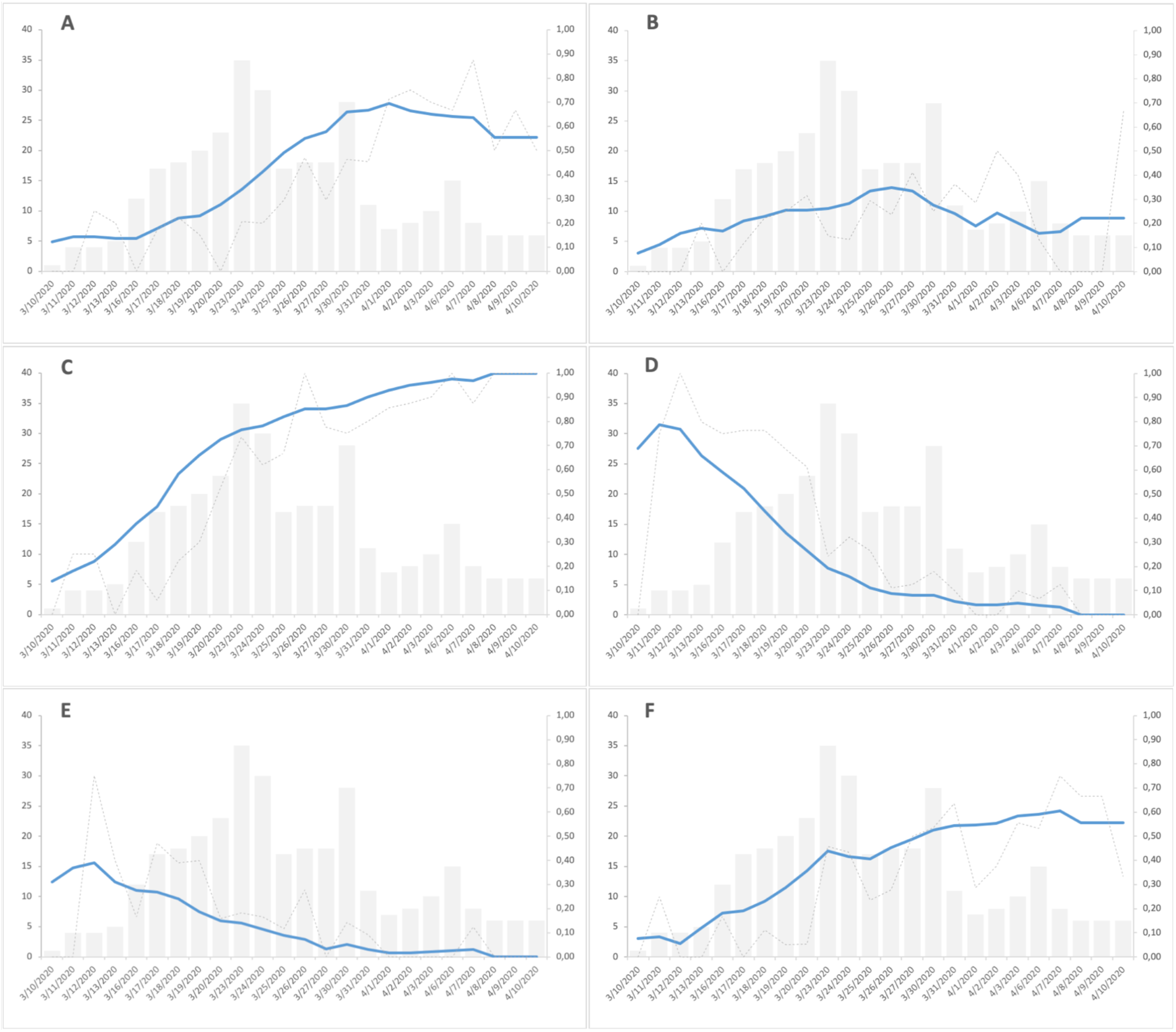
Evolution of exposures reported by positive hospital personnel over time. Grey bars represent the number of COVID-19 cases among hospital staff in the two settings (left y-axis). The grey line represents the crude daily proportion of positive HCW reporting the exposure and the blue line its 7-days mobile mean (right y-axis). A: HCW reporting close contact with a COVID-19 patient with PPE; B: HCW reporting close contact with a COVID-19 patient without PPE; C: HCW reporting wearing mask most of the time or always at hospital; D: HCW reporting close contact with colleagues without masks for more than one hour per day; E: HCW reporting childcare out of the household; F: HCW reporting wearing mask out of hospital.

## Discussion

This prospective study, implemented in real-time in two tertiary hospital settings (adult and pediatric) in central Paris, gives new insights on the dynamics and the sources of exposure of SARS-CoV-2 infections in HCW. Despite a significant number of cases reported, very few had severe clinical presentations. The outbreak in HCW preceded the community epidemic by about one week, and most cases were reported at the very early phase of the French epidemic, before implementation of collective mitigation measures. Control measures (universal masking and PPE for direct care to COVID-19 patients) were rapidly adopted in both hospitals, leading to a decrease of new COVID-19 infections in HCW by late March 2020, while at the same time, the epidemics was still progressing in the community. A residual number of contaminations was reported as of April 1^st^ among HCW, mainly driven by contacts with undiagnosed patients or colleagues without protection. Conversely, children attending community childcare facilities seemed to play a minor role in residual transmissions to HCW that were observed during the second phase of the epidemic.

Compared to previous studies in China (1), the US (5) and the UK (3,4) on SARS-CoV-2 infections in HCW, we did a systematic investigation among all staff members infected with COVID-19 and were able to confront an adult and a pediatric setting. Epidemic curve among HCW was also compared to hospitalization needs in the general population. Therefore, our data collected from the early beginning of the French epidemics give an unprecedentedly reported landscape of HCW infection with COVID-19. Hospital workers were infected regardless of their profession category and their unit of work, suggesting that manifold determinants of transmission are involved. Only three participants had severe forms of COVID-19 and no death were deplored to date. Symptoms were consistent with literature, a majority of cases reporting headaches, fever, cough, asthenia, rhinorrhea, muscle pains and loss of smell or taste.

Preventive measures with implementation of universal masking, reinforcement of hands hygiene and social distancing were applied from March 16^th^ in our two settings. Seven days later, the epidemic curve flattened, although HCW were increasingly exposed to COVID-19 patients and the outbreak was concomitantly at its peak in the region Ile-de-France. This shift is consistent with the SARS-CoV-2 incubation interval (7) and data reported elsewhere (3). Our data suggest that when IPC measures are correctly applied, working in a COVID-19 unit does not appear to ffavorCOVID-19 infection. In particular, our results support that medical masks for general patient care (out of aerosol-generating procedures) were efficient to protect HCW and prevent contaminations, which is consistent with most current guidelines (10,13). This observation provides additional evidence that SARS-Cov-2 is primarily transmitted via droplets, despite discussion in the scientific literature on the possibility of airborne transmission, raised by experimental studies led in laboratory conditions (8). Our results support the key role of universal masking and hand hygiene to offer protection. Lack of compliance of mask wearing reported by positive HCW during breaks, together with recent data evidencing viral shedding occurring before symptoms onset (7), also underscore the crucial importance of maintaining physical distancing rules between hospital employees, even if asymptomatic, during out-of-duty activities.

The attack rate was significantly lower in the pediatric hospital. More COVID-19 patients were admitted in the Adult setting, but more importantly, compliance with PPE was higher in the Children setting. One explanation might be the higher awareness and compliance to IPC measures in pediatric setting where gowns and droplet precautions with medical masks are routinely recommended even out of a pandemic context (15). Data obtained by others also suggest that compliance to hands hygiene is better in pediatric wards than in adult’s ones (16).

The role of children in the current pandemic is still widely questioned (17). Children have been initially suspected to play a central role in transmission of SARS-CoV-2. However, recent data have been more balanced and their role remains unclear (18). Although early Chinese data suggested that children were at a similar risk of infection to general population (19), others showed that attack rates in children may be lower than in adults (20,21). Moreover, in a well-documented cluster of 12 COVID-19 cases in French Alps, one child was involved. No COVID-19 secondary case from him emerged, although he attended three different schools and a ski class while symptomatic and whereas other respiratory viruses like picornaviruses or influenzae viruses spread within his contacts (22). Due to the lockdown, majority of schools and childcare facilities were closed since March 12^th^, but few remained open to welcome HCWs’ children in small groups. This choice raised questions on the risk of maintaining circulation of the virus among children of HCW, thus exposing them to the infection. Interestingly in our cohort, the proportion of COVID-19 positive hospital personnel reporting having a child in an out-of-home care service dramatically decreased all over the epidemic period. Thus, residual contaminations of HCW observed at the late phase of the epidemic were unlikely attributable to keeping schools and childcare services open for HCW. This suggests that in case of second wave of the COVID-19 outbreak, keeping children services available for HCW would be acceptable to help asymptomatic hospital staff stay at work. However, access to these collective structures were restricted to HCW and limited to a very small number of children, limiting contacts and therefore the risks of transmissions. Thus, this does not presume on the impact of wide reopening of schools and nurseries after the lift of containment measures.

The main limitation of our study is the lack of a control group (i.e. inclusion of HCW that were not infected by SARS-Cov-2) to formally compare exposures and assess their respective role in transmission. Identification of negative controls is nevertheless difficult. Indeed, sensitivity of the rt-PCR on nasopharyngeal swabs is imperfect (23), therefore identification of controls only based on a negative rt-PCR result is questionable. The gold standard to rule out the diagnosis of COVID-19 is serologic assessment. Performances of these technics are still under investigation in France before large implementation among HCW. We chose to rapidly communicate timely data in order to guide decisions in view of the soon upcoming lift of containment measures. Further investigations are on the way to identify negative controls by serologic assessment and formally compare their exposures to HCW of our cohort. Another limitation is the recall bias which is inherent to the use of questionnaires in epidemiological studies, however infected HCW were interrogated prospectively and shortly after PCR assessment.

Several conclusions can be drawn from our results: (i) HCW are exposed to emerging viral diseases, particularly at the early phase of the epidemic, as illustrated by the overall attack rate of 2.8%; (ii) compliance to control measures increased over the study period, concomitantly with containment of the outbreak among hospital staff; (iii) incidence was lower in HCW of the Children setting, likely related to a better adherence to IPC measures by the pediatric staff and (iv) residual transmissions observed at the late phase of the epidemic among HCW were related to persistent exposures with undiagnosed patients or colleagues and not to contacts with children attending out-of-home care facilities.

## Data Availability

Authors declare that all data referred in the manuscript is fully available.

## Authors’ Contributions

AC, JL, OL and SK designed the study and drafted the paper. AC, JL, ML, FR, OL and SK contributed to data analysis and interpretation. All authors critically revised the manuscript for important intellectual content and gave final approval for the version to be published. OL and SK had full access to all the data in the study and had final responsibility for the decision to submit for publication.

## Declaration of interests

AC reports personal fees and non-financial support from Janssen Cilag, outside the submitted work. RG reports personal fees and other from Sanofi, personal fees from MSD, personal fees from Eumedica, personal fees from Pfizer, personal fees from Frezenius, personal fees from Cubist, personal fees from Correvio, personal fees from Astellas, outside the submitted work. PF reports personal fees and non-financial support from MSD France, personal fees from ViiV Healthcare, non-financial support from Gilead Science, personal fees and non-financial support from Janssen Cilag, personal fees from Medtronic SAS, non-financial support from Astellas, non-financial support from GSK, outside the submitted work. MLV reports non-financial support from Biomerieux, non-financial support from Diasorin, outside the submitted work. OL reports personal fees from Pfizer, personal fees from Astellas, personal fees from MSD, personal fees from Gilead, outside the submitted work. SK reports research grants from bioMérieux, personal fees from Accelerate Diagnostics, bioMérieux, MSD and Menarini, and travel expenses from bioMérieux, Astellas, MSD and Pfizer, outside the submitted work. All other authors declare no conflict of interests.

## Acknowledgments

The authors warmly thank medical students involved in data collection: Laurence Clastres, Mathilde Lehmann, Aline Pellegrini, Marine Sisouvan, Ilana Slotine, Diem Soubou and Abigaëlle Vergnet, the following physicians and nurses who actively contributed to the screening: Claire Aguilar, Chantal Delmas, Claudine Duvivier, Muriel Fortier, Béatrice Grandordy, Marc Lecuit, Léonie Meyer,

Gabrielle Paluszek, Guillemette Thin and Christine Vinter, as well as Jeanne Marty for providing hospital data and Bruno Coignard for his help on data interpretation.

